# Design and validation of a clinical whole genome sequencing-based assay for patient screening in a large healthcare system

**DOI:** 10.1101/2025.07.16.25331598

**Authors:** Josiah T. Wagner, John T. Welle, Isabelle A. Lucas Beckett, Kate R. Emery, Benjamin A. Cosgrove, Krzysztof Olszewski, Nick Wagner, Tucker C. Bower, Li Chi Yuan, Eric M. Shull, Kathleen Jade, Jon Clemens, Andrew T. Magis, Mary B. Campbell, Ora K. Gordon, Carlo B. Bifulco, Brian D. Piening

## Abstract

Population genetic screening is rapidly emerging as a key methodology in the clinical laboratory for detecting actionable genomic conditions. While current clinical methods are largely focused on targeted gene panels, the increasing efficiency of next-generation sequencing (NGS) platforms permits the use of whole genome sequencing (WGS) for routine clinical applications. The key advantage of WGS is that the complete genome produced by a single sequencing event can form the basis for a patient’s genomic health care record for reanalysis throughout a patient’s lifetime. Here, we developed a scalable clinical WGS-based lab developed procedure (LDP) for heritable disease gene testing and pharmacogenomics (PGx). We performed extensive validation across a range of blood, saliva, and reference specimens. The clinical deliverable for the WGS LDP was 78 genes associated with actionable genomic conditions and 4 PGx genes. The validation cohort consisted of samples from 188 study participants that were orthogonally sequenced at commercial reference laboratories and additional reference materials. The deployed LDP was then used to sequence over 2000 patients as part of a broader clinical implementation study (“Geno4ME”). We demonstrate that the WGS LDP has excellent sensitivity, specificity, and accuracy, thus supporting WGS as a viable method for broad clinical screening.

## Introduction

Identification of individuals at risk for hereditable genetic conditions or suboptimal drug dosing provides opportunities for life-saving medical intervention and improvement in treatment outcomes. Accumulating population genomics evidence suggests that a significant number of individuals carry a clinically actionable genetic variant that has risk of disease, yet are unaware of their genetic risks until symptomatic [1–4]. Additionally, recent insights suggest that up to 98-99% of individuals have one or more genetic variants that can impact drug efficacy and safety [5]. Pharmacogenomics (PGx), which focuses on the identification of genomic variants in an individual that modulate pharmacokinetics and pharmacodynamics of specific classes of drugs, can have direct implications for prescribing guidelines [6]. The high prevalence of individuals with actionable genomic variants has resulted in growing interest in genomic screening tools for broad population health applications [7]. However, there remains a need for genomic screening tools with gene panels that can be rapidly expanded, quickly interpreted, and with a scalable workflow.

A comprehensive genomic assay evaluable for both heritable genomic conditions and PGx in populations should ideally cover a large and diverse panel of gene loci. While most modern genomic screening tools have been effective at characterizing small genomic variants (approximately less than 50 bp in size), extensive evidence suggests that large copy number variants in the human population are a contributor to heritable disorders [8]. Thus, an ideal procedure should be able to characterize multiple variant types, including single-nucleotide variants (SNVs), multi-nucleotide variants/polymorphisms (MNVs), insertions, deletions, and copy-number variants (CNVs). High-throughput methods for simultaneously characterizing single nucleotide variants in multiple genes, such as MALDI-TOF and SNV array-based methods, have been applied in a population health context [9–11]. To characterize larger variants, microarray-based comparative genomic hybridization (array CGH or aCGH) has been applied clinically [12–14]. Characterization of diverse variant types using these methods can require multiple assays to acquire a comprehensive genomic profile and often there is difficulty with resolving novel and/or complex variants. To address these limitations, whole exome sequencing (WES) and whole-genome sequencing (WGS) have been applied in a variety of population genomic health applications, such as healthy population screening [15–18], unselected research cohort screening [1, 19–24], and newborn screening [25–27]. Both WES and WGS allow for single-nucleotide resolution of variants and have been demonstrated to capture a significant number of gene- and exon-level CNVs with potential clinical importance [28, 29]. WGS, when compared to WES, has reduced variant allele capture bias, can potentially profile more complex chromosomal rearrangements, and can be expanded to noncoding and intergenic regions [30]. Recent advances in PCR-free WGS have led to increased variant detection sensitivity and retention of complex genotypes (e.g. repetitive regions) when compared to conventional WGS [31, 32]. Therefore, a PCR-free WGS-based assay is ideal for comprehensively evaluating variation in coding regions while providing opportunity for future region expansion as additional loci of interest become evident.

Although using a WGS-based assay as a primary method of diagnosis holds significant promise as a tool for rare diseases and large-scale population health genomics [19, 33], few studies have evaluated the practicality and performance of WGS for returning clinically-actionable results in a healthcare setting. Here, we developed a clinical PCR-free WGS-based lab developed procedure (LDP) for hereditable disease testing and PGx. Using a large cohort, we validated our WGS-based assay for variant detection, variant pathogenicity classification, and PGx interpretation against orthogonal panel testing at outside reference laboratories. Additionally, we determined if DNA originating from either blood or saliva specimens impacted WGS assay performance. Our results support the feasibility and high diagnostic accuracy of using a WGS-based assay as a core population health tool for evaluating clinically actionable genomic variants.

## Methods

### Participant selection and sample collection

This study was reviewed and approved by the Providence Institutional Review Board (approval number STUDY2020000637). The validation of the WGS LDP was performed in support of the “Genomic Medicine for Everyone” (Geno4ME) clinical implementation study, involving WGS of Providence patients, return of selected clinical results, and genome banking for future research [34]. Study participants were patients in the Providence system and informed consent was obtained from all participants using an in-house developed automated electronic consent platform. Information on participants’ personal/family history of cancer and/or cardiovascular conditions, ethnicity, and current medications taken was obtained using a self-reported survey. Whole blood was collected using venipuncture and stabilized in standard clinical use EDTA tubes. Saliva was collected and stored using DNA Genotek Oragene-DNA saliva DNA collection kit (DNA Genotek #OGR-600). In total, 120 whole blood and 70 saliva specimens were collected in the WGS Validation cohort, with 60 participants providing paired whole blood and saliva samples for cross-sample validation (N = 189 unique participants).

### Genomic condition gene selection for analysis

For selected genomic conditions, we evaluated variant pathogenicity in 78 genes determined to be clinically actionable and reportable by the ACMG as secondary findings in clinical exome and genome sequencing and/or National Comprehensive Cancer Network (NCCN) guidelines [35, 36]. The complete list of genes and genomic conditions analyzed in this study are shown in Figure 1.

**Figure 1.**
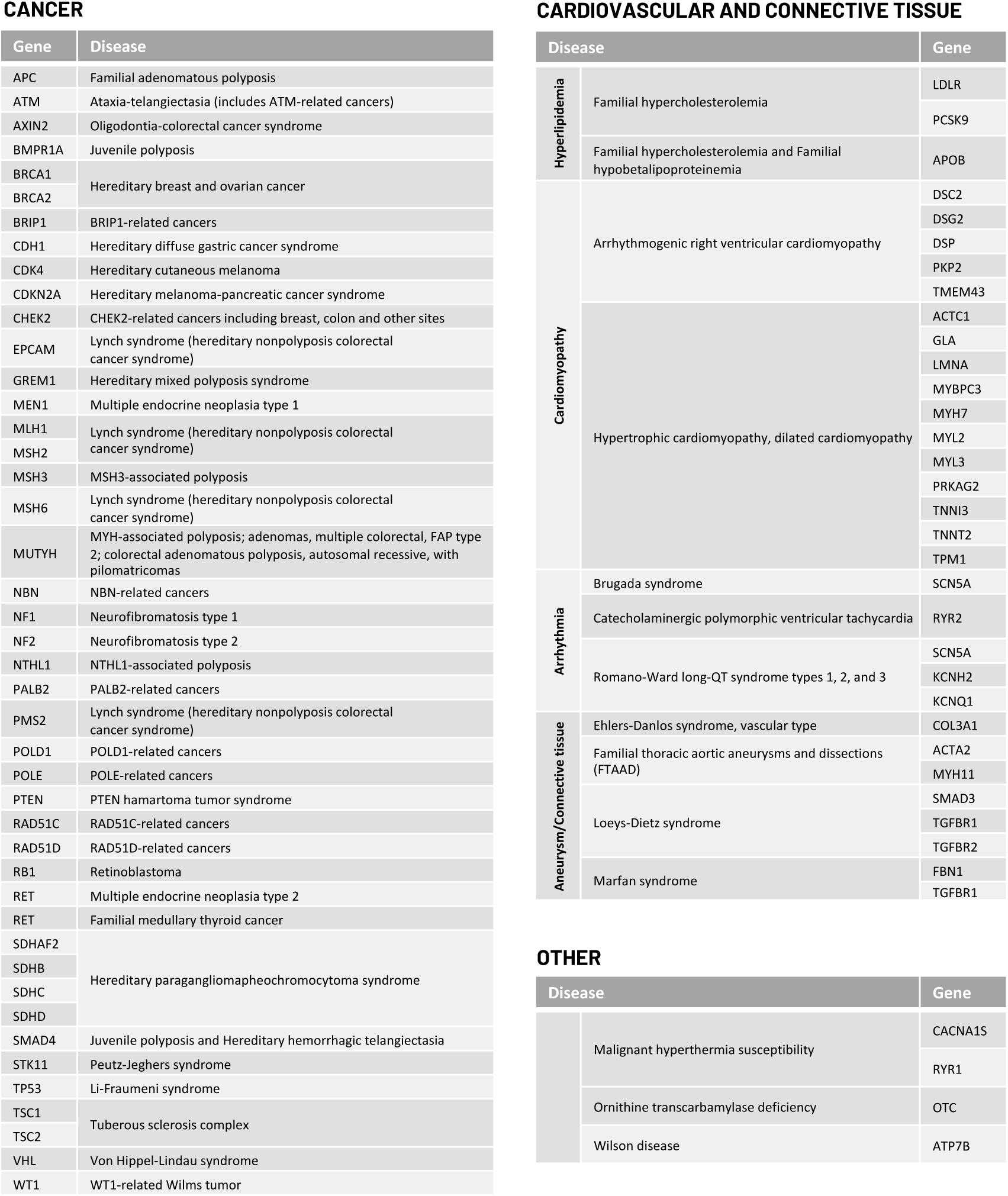
Genes tested for the Geno4ME-LDP heritable disorder panel and their associated diseases.

### Sample genomic DNA extraction, sequencing, QC, and variant calling

Genomic DNA for whole genome sequencing was extracted using the Qiagen QIAsymphony DSP Midi Kit (catalog 937255). Whole genome next-generation sequencing (NGS) libraries were prepared from 300 to 500 ng gDNA with the Illumina DNA PCR-Free Prep, Tagmentation kit (catalog 20041795). Sequencing was performed on the Illumina NovaSeq 6000 with 24 libraries loaded per S4 flow cell and a target depth of 30X coverage. As a sequencing quality control, the Illumina PhiX Control v3 Library was sequenced on every WGS with error rates of less than 1% considered passing. In addition, every 100 samples a germline variant quality control sample containing known germline DNA variants was extracted, sequenced, and annotated in the same manner as other patient samples. The germline variant quality control was prepared as a WGS library, sequenced, and annotated in the same manner as patient samples. Intra-run variation (within-run) was determined by preparing and sequencing three WGS replicates of sample gme-wes-25 and sequencing the libraries on the same WGS run. Inter-run (between-run) variation was determined by preparing and sequencing three WGS replicates of DNA from control sample NA12878 and sequencing across three separate WGS sequencing runs. Variant calling and pharmacogenetic genotyping were performed using standard analysis pipelines on the Illumina DRAGEN (Dynamic Read Analysis for GENomics) Bio-

IT Platform (v3.9.5) with the following flags: --enable-map-align true, --enable-map-align-output true, --enable-duplicate-marking true, --enable-sort true, --vc-combine-phased-variants- distance 3, --vc-enable-roh true, --vc-enable-baf true, --vc-enable-phasing true, --enable-variant-caller true, --enable-cnv true, --cnv-enable-self-normalization true, --cnv-enable-tracks true, --vc-enable-roh true, --vc-enable-baf true, --vc-enable-phasing true, --enable-variant-caller true, --enable-cnv true, --cnv-enable-self-normalization true ,--cnv-enable-tracks true, --cnv- segmentation-mode slm, --cnv-interval-width 250, --enable-sv true. Variants with VAF less than 10% were excluded from further analysis. Regions for gene variant analysis were extracted using coordinates on the hg19 (GRCh37) reference derived from the longest Refseq transcript [37, 38]. These regions for analysis were extended by an additional 1,000 bases upstream and downstream of the first (5’ UTR) and last (3’ UTR) exons, respectively. Exon-level and whole gene-level CNV and structural variation (SV) was detected using the DRAGEN and/or Manta CNV callers included as part of the Illumina DRAGEN Bio-IT Platform. Coverage metrics were calculated using deepTools2 [39]. Reads not mapping to hg19 were discarded from further analysis.

### Validation of WGS variant calling method to variants from patient electronic medical records

Initial validation of the WGS DRAGEN variant calling pipeline was performed by comparison of WGS DRAGEN variants to clinically significant genomic variants obtained from patient electronic health records (EHRs). Genomic DNA for WGS was extracted, sequenced, and analyzed as described above for 30 bio-banked patient blood samples collected under IRB protocol STUDY2018000254, an internally funded Providence protocol of germline P/LP carriers of hereditary cancer risk. The Providence Genetics research program utilizes the Progeny database to curate and maintain highly annotated screening, disease and genetic testing data for participants in the biorepository. Sample selection was drawn from the Progeny database of de-identified samples. Participants provided DNA samples for future use in discovery and all available EHR genomic testing information (EHR Comparison validation group). Samples were selected based on variety of genes to be within the Geno4ME suite broad variant profile (truncating, missense, indel and genomic rearrangements, with available EHR genomic testing information (EHR Comparison validation group). A positive variant match was determined by the presence of an expected gene coding and/or protein change as described in the patient EHR to a variant in the WGS DRAGEN pipeline that passed all quality filters.

### Classification of variants

To classify variants in genes with an associated genetic condition, pathogenicity was assessed based on criteria from the joint American College of Medical Genetics and Association for Molecular Pathology Standards and Guidelines (ACMG/AMP criteria from 2015) [40]. PP5 or BP6 criteria were not considered for final variant pathogenicity classification as recommended by Biesecker *et al*. [41]. Prior to manual variant curation, variants were initially classified using the Artificial Intelligence Classification Engine (ACE), an automated ACMG classification algorithm in the Fabric Enterprise platform (Version 6.18.X) [42]. All variants, including CNVs and SVs, were uploaded to the Fabric Enterprise platform for automated interpretation prior to manual curation or interpretation. Variants were targeted for manual curation if initially classified as pathogenic or likely pathogenic (P/LP) by ACE and/or the interpretation for the associated genetic condition in the ClinVar genomic annotation aggregation database was P, LP, conflicting, or not provided [43]. Evaluation of variant population frequency, *in silico* predictions of variant effect, and statistical support for pathogenicity using the ACMG/AMP criteria were performed using tools in the Fabric Enterprise platform. Literature review for variants identified for manual curation was assisted by Mastermind Genomic Search Engine [44]. For putative variants of uncertain significance (VUS) and P/LP variants, alignment quality of the region was manually inspected using Integrative Genomics Viewer (IGV) for read and alignment quality [45]. The overall workflow for WGS and interpretation we termed the Geno4ME LDP as is henceforth referred to below. The Geno4ME LDP method was used for comparison to all reference methods (Figure 2).

**Figure 2.**
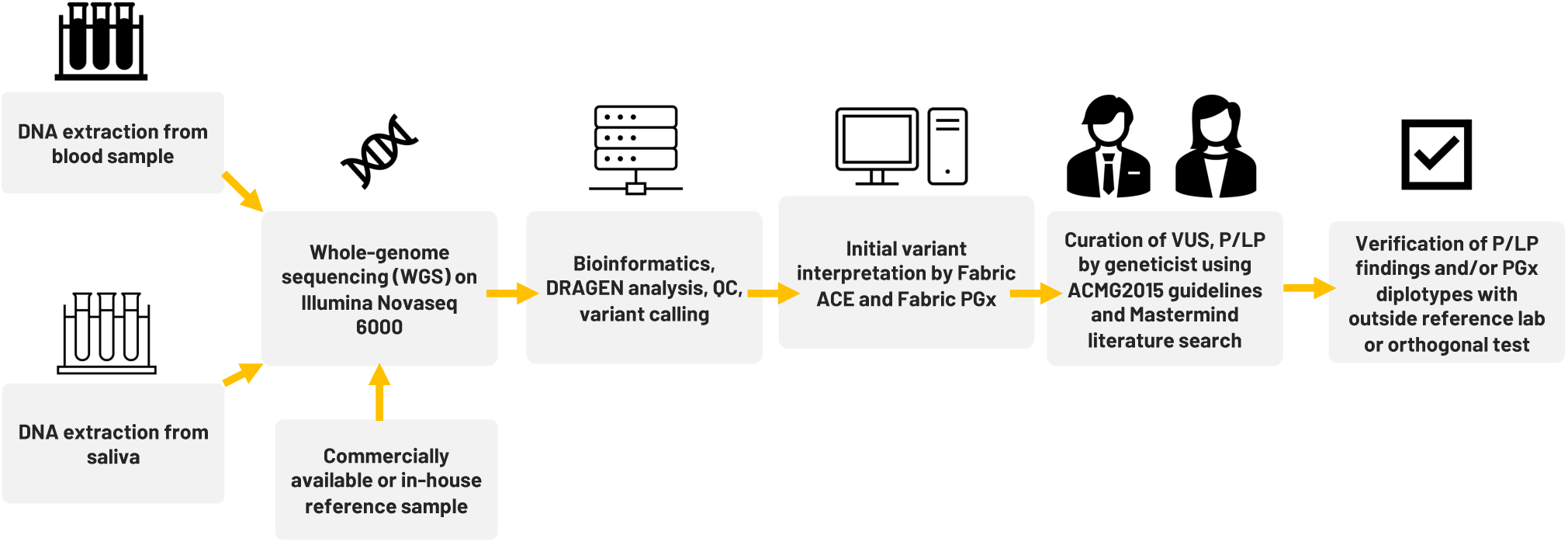
Overview of workflow for WGS sample processing, interpretation, and comparison to outside references for assay validation. This complete workflow is referred to as Geno4ME LDP.

### Geno4ME LDP orthogonal validation with outside reference methodologies

Sensitivity, specificity, and accuracy of the Geno4ME LDP for variant calling was determined by comparison to a CLIA-certified commercial molecular laboratory using an outside reference method (OS-ORM). The OS-ORM was based on hybrid-capture NGS for all 78 genes associated with a genomic condition surveyed in this study were covered by OS-ORM Panel A, OS-ORM Panel B, or both (Supplemental Table S1). Because not all VUS were reported by the outside provider due to differences in outside provider classification or ORM panel return of results (RoR) criteria, only variants identified by the OS-ORM to be P/LP were considered to be true positives for comparison. For genes where a different transcript was selected between the Geno4ME LDP and ORM, gene variants were remapped to the Geno4ME LDP selected transcript (Supplemental Table S2). In total, Geno4ME LDP variant calling results for 188 samples (119 whole blood and 69 saliva from the WGS Validation ORM group, Supplemental Spreadsheet A) were validated by comparison the OS-ORM. Sensitivity, specificity, and accuracy of the Geno4ME LDP for variant calling was further tested by comparison to germline data from a previously-described cohort of 25 cancer patient reference samples sequenced by a clinically-validated WES assay (WES Comparison Validation Group, Supplemental Spreadsheet A) [46].

Reference DNA samples for CNV caller validation Geno4ME LDP were obtained from the National Institute for Biological Standards and Control (NIBSC, UK Stem Cell Bank Blanche Lane South Mimms Potters Bar Herts. EN6 3QG, NIBSC code: 11/218-XXX). Seven purified human genomic DNA samples with or without CNV variants in MLH1 and MSH2 were tested using the Geno4ME LDP and compared to the known copy number genotypes provided by the manufacturer (Table 1). Accuracy of the Geno4ME LDP was measured by identification of expected exon CNVs alterations characterized by the manufacturer.

**Table 1.** Expected SV/CNV in control NIBSC samples tested using the Geno4ME LDP method.

| Sample ID | Expected SV/CNV |
| --- | --- |
| NIBSC 1 | None |
| NIBSC 2 | None |
| NIBSC 3 | *MSH2* deletion exons 1-6, heterozygous |
| NIBSC 4 | *MSH2* deletion exon 7, heterozygous |
| NIBSC 5 | *MSH2* deletion exons 1-2, heterozygous |
| NIBSC 6 | *MSH2* deletion exon 1, heterozygous |
| NIBSC 7 | *MLH1* exon 13 amplification (three or more copies) |

### Geno4ME LDP PGx genotyping and PGx phenotyping

Five PGx genes with gene-drug prescribing guidelines were selected based on the published joint recommendations from Clinical Pharmacogenetics Implementation Consortium (CPIC) and the U.S. Food and Drug Administration [47–51]. For these five PGx genes, variants were selected based on recommendations from the Association for Molecular Pathology (AMP), the College of American Pathologists (CAP), and/or CPIC [51, 52]. The pre-selected defining PGx variants, the logic to define genotypes as well as the genotypes to phenotype mapping were implemented into Fabric Genomics cloud platform and used as part of the Geno4ME LDP (henceforth referred to as Geno4ME LDP PGx). The PGx panel considered for validation included 7 gene-drug pairs that were selected based on FDA and CPIC guidelines (Table 2). For *CYP2C19*, both Tier 1 (*2, *3, and *17) and Tier 2 (*4A, *4B, *5, *6, *7, *9, *10, and *35) alleles were included per AMP/CAP recommendation, *CYP2C9* Tier 1 alleles (*2, *3, *5, *6, *8, and *11), *VKORC1* (c.-1639G>A, rs9923231), *CYP4F2* (*3), and the single variant rs12777823 (*CYP2C* cluster) were included as recommended in the CPIC guideline for warfarin. The genotype to phenotype mapping was based on PharmgKB, CPIC, and PharmVar annotations (Supplemental Table S3) [51, 53, 54]. *CYP2C19*, *CYP2C9*, and *CYP4F2* alleles negative for assayed variants were designated as *1.

**Table 2.** List of drugs and their associated PGx genes assayed.

| Drug(s) | Drug type | Associated gene(s) |
| --- | --- | --- |
| Warfarin | Anticoagulant/antiplatelet | *VKORC1, CYP2C9, CYP4F2,*<br>*rs12777823* |
| Clopidogrel | Anticoagulant/antiplatelet | *CYP2C19* |
| Lansoprazole<br>Omeprazole<br>Pantoprazole | Proton pump inhibitors | *CYP2C19* |
| Citalopram<br>Escitalopram | Selective serotonin reuptake inhibitors | *CYP2C19* |

### Validation of Geno4ME LDP PGx genotyping

Validation of *CYP2C19*, *CYP2C9*, *CYP4F2, VKORC1* genotyping by Geno4ME LDP PGx was performed by comparison to a CLIA-certified commercial molecular laboratory using an outside reference method for PGx (OS-ORM PGx). The outside reference method for PGx validation was MassARRAY genotyping (Invitae). Accuracy of Geno4ME LDP PGx genotyping against the ORM PGx was performed using the same 188 samples used for validating the Geno4ME LDP variant call concordance (WGS Validation ORM group, Supplemental Spreadsheet A). In addition, accuracy of Geno4ME LDP PGx genotyping was further validated by comparing Geno4ME LDP PGx to 18 previously characterized cell lines/DNA samples (WGS Validation PGx validation group). The following cell lines/DNA samples were obtained from the NHGRI Sample Repository for Human Genetic Research at the Coriell Institute for Medical Research: NA7019, NA7029 NA07439, NA10847, NA12717 NA17641, NA18524, NA19109, NA23275, NA24008, HG00436, HG00589, HG01190, NA07348, NA12003, NA12878, NA19207, and NA19785. These cell lines/DNA samples were used to validate the Geno4ME LDP PGx for *CYP2C19, CYP2C9, CYP4F2*, and *VKORC1* diplotypes based on previously described diplotypes [55]. Genotypes not supported by the Geno4ME LDP PGx were not evaluated for accuracy when comparing to the NHGRI Coriell samples. The single variant rs12777823 was not supported by the OS-ORM PGx or NHGRI Coriell reference method and was therefore not compared to the Geno4ME LDP PGx method.

### Statistics and plots

Sensitivity, specificity, Positive Predictive Value (PPV), Negative Predicative Value (NPV), and 95% confidence intervals (Cis) were calculated using the MedCalc Diagnostic Test evaluation calculator online tool (https://www.medcalc.org/calc/diagnostic_test.php). Graphs were created using Graphpad Prism (v10.1.1) or Rstudio (v2023.06.01, R version 4.3.1). For comparisons of the Geno4ME LDP for gene variant calling to the ORM and WES-RM, a positive gene match was determined if all variants detected within a gene matched between the Geno4ME LDP and reference method. Negative genes were determined if no variants were reported in either the Geno4ME LDP or reference method. Due to assay limitations, MSH3 exon 1 and the pseudogene region of PMS2 were excluded for comparisons of the Geno4ME LDP to the OS-ORM and WES-RM. A summary of validation group sample sizes and their purposes is shown in Table 3.

**Table 3.** Summary of samples used for validation in this work.

| Validation Group | N<br>Individuals | N<br>samples | Purpose |
| --- | --- | --- | --- |
| EHR Comparison | 30 | 30 | Validation of DRAGEN WGS variant calling |
| WGS Validation ORM | 188 | 188 | Validation of Geno4ME LDP variant calling and PGx diplotyping to OS-ORM |
| WGS Validation SB <sup>a</sup> | 60 | 120 | Validation of Geno4ME LDP variant calling and PGx diplotyping between Saliva and Blood samples |
| WGS Validation CNV | N/A | 7 | Validation of CNV calling using the Geno4ME LDP to previously characterized DNA samples |
| WES Comparison | 24 | 24 | Validation of Geno4ME LDP variant calling to orthogonal WES-ORM |
| WGS Validation PGx | N/A | 18 | Validation of Geno4ME LDP PGx using previously characterized Coriell cell lines |

<sup>a</sup>59 samples for the Saliva-Blood comparison overlap with the WGS Validation ORM validation group

## Results

### Cohort composition and sequencing characteristics

For the WGS Validation ORM and WGS Validation SB validation groups the average age at time of collection was 56.5 years (SD = 13 years, N=189 individuals). Out of these 189 participants, 157 self-reported sex as female and 32 as male. 60 participants provided matched saliva and blood samples. The average WGS sequencing coverage over human genome, separated by validation group, is shown in Table 4. Median coverage over genome per group ranged from 35.2 – 41.9X median per-base coverage. Samples used for inter-run variation analysis had a mean coverage over genome of 34.2X (SD = 1.7, CV = 4.9%). Samples used for intra-run variation analysis had a mean coverage over genome of 40.9X (SD = 8.8, CV = 21.4%). Full sequencing metrics for individual samples and by validation group are shown in Supplementary Figures S1-S3 and Supplemental Spreadsheet A.

**Table 4.**
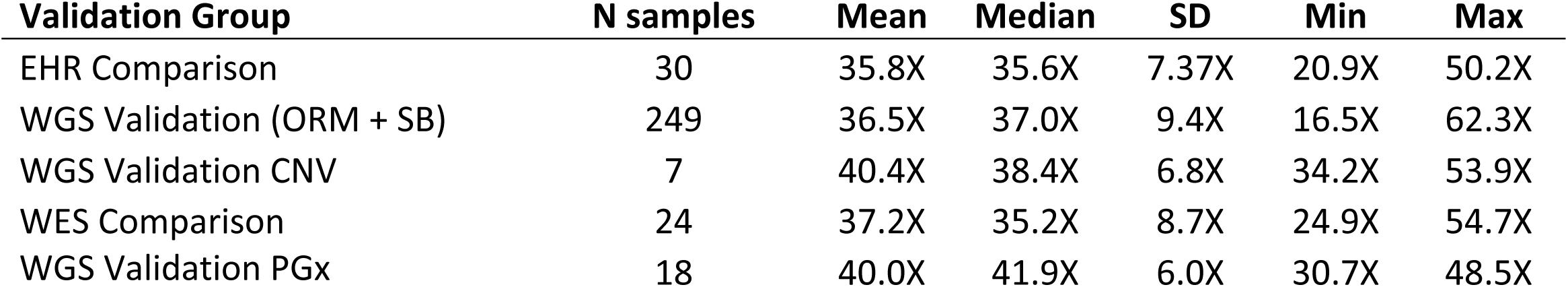
Coverage over genome metrics calculated by validation group.

### Concordance of DRAGEN WGS variant calling method to variants from patient EHRs

A total of 33 variants identified across 30 patient EHRs were used for comparison to the WGS DRAGEN variant caller (Supplemental Spreadsheet B). Most identified variants were missense (Figure 3A) and the most frequent gene represented was *BRCA1* (Figure 3B).

**Figure 3.**
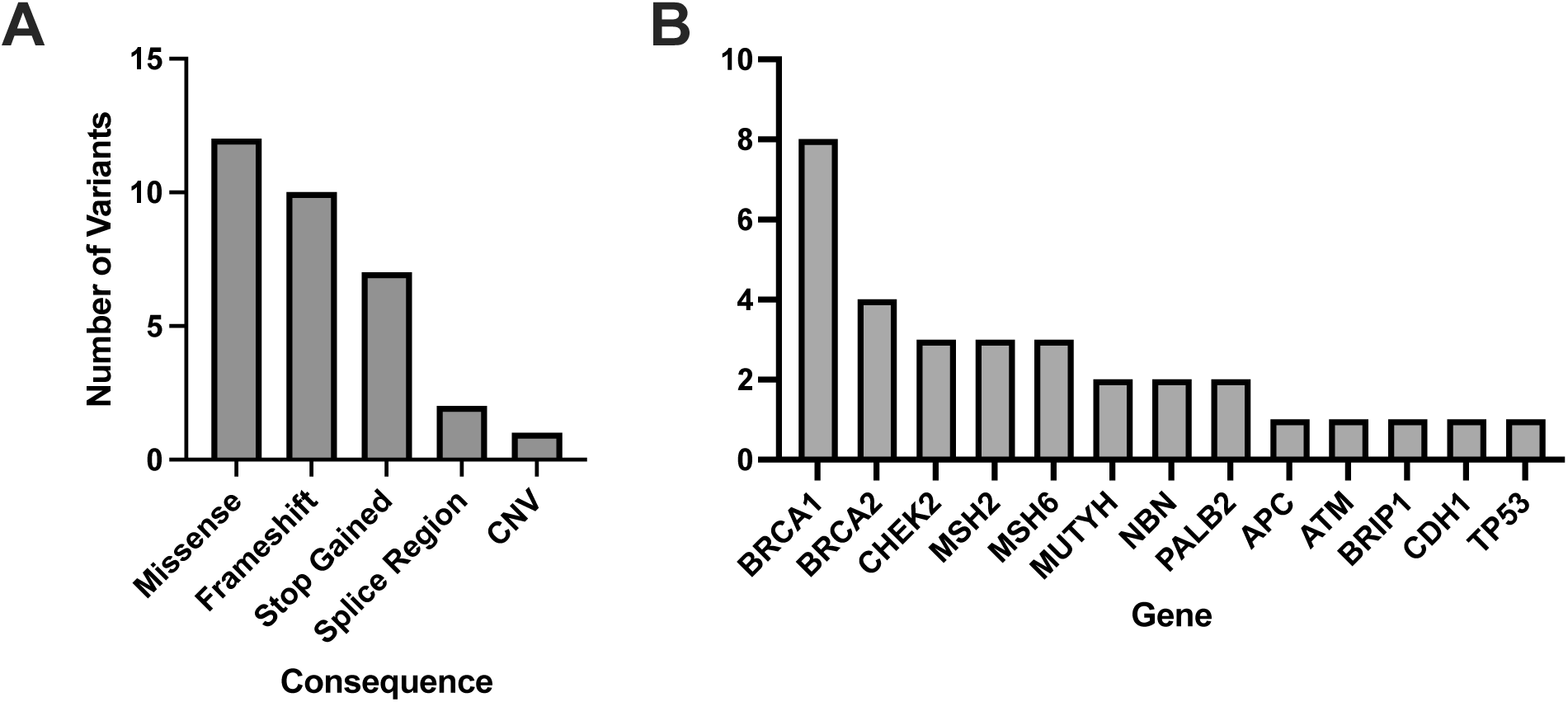
Molecular consequences of all matched variants identified in patient EHRs and compared to the DRAGEN WGS variant calling. A total of 33 variants across 30 patient EHRs had variant matches in the DRAGEN WGS variant calling pipeline and the majority were missense (A). Out of the variants identified in patient EHRs, 13 genes were represented (B).

Patient HR23 was found to have a large, multi-exon heterozygous *BRCA1* deletion identified by DRAGEN (*BRCA1* c.678_4740del, del *BRCA1* exons 10-15/24) and this deletion had 99% overlap with the deletion described by EHR (*BRCA1* c.671_4675del). Overall, all 33 variants identified in patient EHRs were identified using the WGS DRAGEN variant caller.

### Concordance of Geno4ME LDP variant calling with the ORM

Within the WGS Validation ORM group, an average of 453.2 total variants per sample (SD = 141.7) were evaluated by the Geno4ME LDP. Most identified variants were SNVs, MNVs, or indels (N=188, Supplementary Figure S4). Using the Geno4ME LDP, a total of 17 genes across 17 different participants were positive with a P/LP variant and most of these P/LP variants were missense (Figure 4A, Supplementary Spreadsheet C). The results of the Geno4ME LDP were compared to the OS-ORM and were found to match with 100% concordance (Table 5, Supplementary Figure S5). Overall, 7 different genes with P/LP variants were represented in the WGS Validation ORM group (Figure 4B).

**Figure 4.**
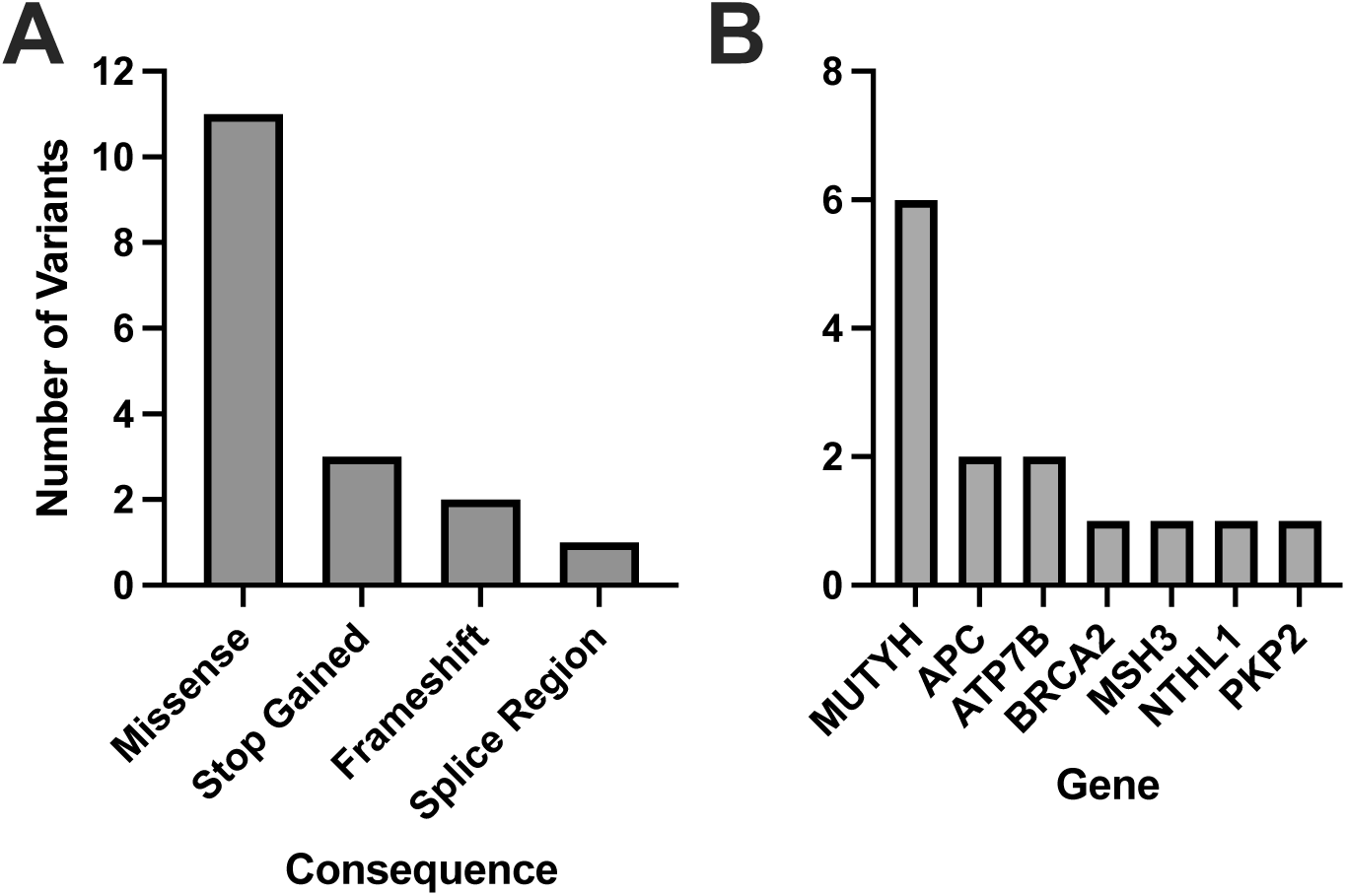
Molecular consequences and distribution of genes with matching P/LP classifications for all matched variants identified in the WGS Validation cohort. The WGS Validation cohort was tested using the Geno4ME-LDP and OS-ORM in parallel. A total of 17 variants across 17 participants had matches to the ORM and the majority were missense (A). Out of 14 P/LP variants that matched between the WGS Validation cohort and the OS-ORM, 7 genes were represented and the most frequent gene with P/LP variants was *MUTYH* (B).

**Table 5.** Concordance of the Geno4ME LDP and the ORM for identifying gene SNVs/indels in the WGS Validation cohort.

|  |  |
| --- | --- |
| **Cohort** | WGS Validation ORM |
| **N samples** | 188 |
| **Experimental Method** | Geno4ME LDP |
| **Reference Method** | Hybridization-based NGS |
| **Positive genes confirmed by Reference Method** | 17/17 |
| **Negative genes confirmed by Reference Method** | 14382/14382 |
| **Effect size** | **Percent (95% CI)** |
| Sensitivity | 100% (80.5% – 100%) |
| Specificity | 100% (100% – 100%) |
| Positive Predictive Value | 100% (80.5% – 100%) |
| Negative Predictive Value | 100% (100% – 100%) |

### Performance of Geno4ME LDP for initial classification of variants

Across the 188 samples in the WGS Validation ORM validation group, 13,096 SNV, MNV, or indel variants were initially evaluated and classified by ACE in the Fabric Enterprise platform (Table 6). Of the 455 variants classified by ACE as VUS, 250 were reclassified as benign following curation. Most of these 250 variants reclassified from VUS to benign consisted of two missense variants that were interpreted by automated pipelines as MNVs without a presence in population databases (dbSNP reference alleles rs386638457 and rs386643884) instead of separate, high population frequency SNVs. Eight variants initially classified by ACE as VUS were reclassified to pathogenic following manual curation. One variant, the in-frame insertion variant *WT1* c.378_392dup p.Ala127_Pro131dup, was classified by ACE as LP but after manual curation was reclassified as VUS. Overall, no variants classified by ACE as B/LB were later found to be LP/P following comparison to variant classification results from the ORM or manual review.

**Table 6.**
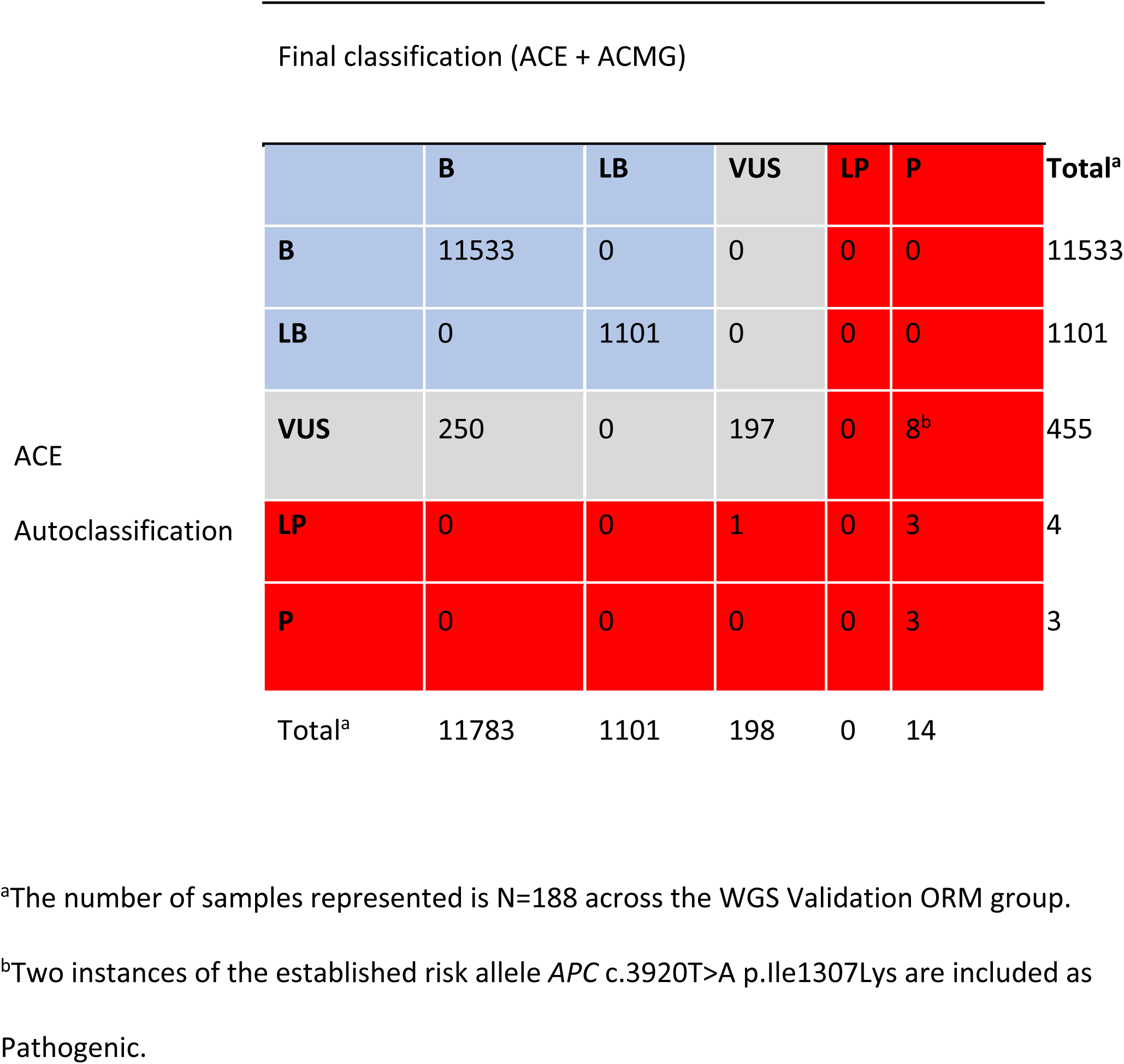
Performance of the ACE autoclassification software for initial classification of variants in the WGS Validation ORM group heritable disorder gene panel.

### Concordance of Geno4ME LDP and WES-RM variant calling

A total of 78 variants across 25 samples were identified using the Geno4ME LDP and/or the WES-ORM within the WES Validation ORM validation group (Table 7, Supplementary Spreadsheet D). Most of the identified variants were missense (Figure 5A). Of the 78 variants identified, 63 unique variants were represented across 26 different genes (Figure 5B). All variants were identified in both the Geno4ME LDP and WES-ORM for each sample, except for a *TP53* c.854A>T p.Glu285Val variant in sample gme-wes-19 that was present at 27% VAF only in the previously sequenced WES data. Investigation of the raw WGS DRAGEN variant calling data for the sample revealed two supporting reads for the *TP53* c.854A>T p.Glu285Val variant (depth = 15) but a low variant quality score (Qual = 5.18) that resulted in filtering by the Fabric Enterprise platform. Review of the patient medical record revealed that a matched solid tumor specimen that was sequenced using hybrid-capture NGS and contained the *TP53* variant at 5% VAF. Using the WES-ORM as the reference method, the Sensitivity and Specificity of Geno4ME LDP for variant calling was 98.72% and 100%, respectively (Table 7, Supplementary Figure S6).

**Figure 5.**
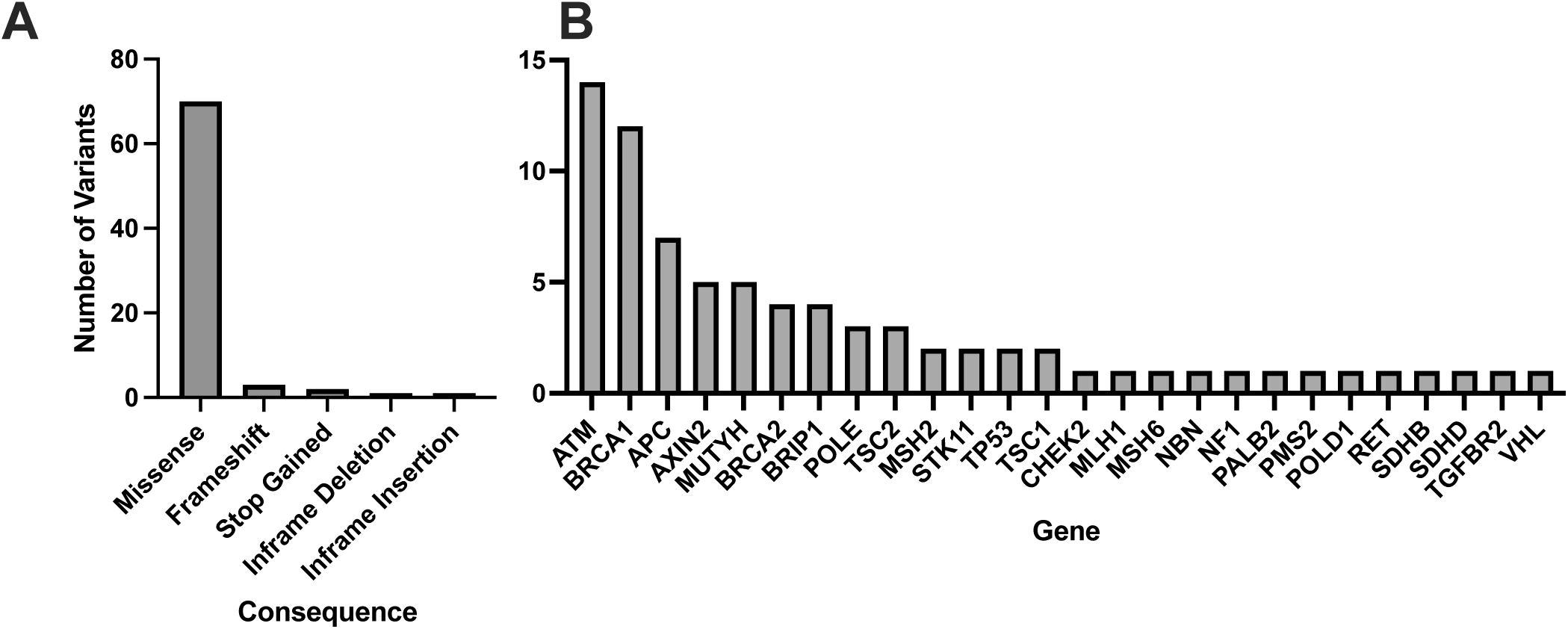
Molecular consequences and genes represented for of all matched variants identified comparing Geno4ME LDP to WES-ORM. A total of 78 variants were identified across all samples, with the majority being missense (A), and 26 unique genes were represented (B).

**Table 7.** Variant concordance between the Geno4ME LDP and WES-ORM in the WES Comparison cohort.

|  |  |
| --- | --- |
| **Cohort** | WES Comparison |
| **N samples** | 25 |
| **Experimental Method** | Geno4ME LDP |
| **Reference Method** | WES |
| **Positive genes confirmed by Reference Method** | 77/78 |
| **Negative genes confirmed by Reference Method** | 874/874 |
| **Effect size** | **Percent (95% CI)** |
| Sensitivity | 98.7% (93% – 100%) |
| Specificity | 100% (99.6% – 100%) |
| Positive Predictive Value | 100% (95.3% – 100%) |
| Negative Predictive Value | 99.9% (99.2% – 100%) |

### Performance of the Geno4ME LDP for identification of large CNVs

Seven control NIBSC samples were tested using the Geno4ME LDP to evaluate CNV caller accuracy at single and multi-exon resolution. All five samples with CNVs characterized by the sample provider were identified using one or both GenoME LDP CNV calling algorithms to affect the same expected gene exons with the same zygosity and CNV type (Supplementary Spreadsheet E). Two samples expected to be negative for CNV alterations were also negative by Geno4ME LDP. Overall, the concordance between the CNV alterations described by the sample provider for the seven control NIBSC samples and the Geno4ME LDP was 100%.

### Concordance of Geno4ME LDP PGx with NHGRI Coriell samples

Eighteen samples obtained from NHGRI Coriell with known *CYP2C19*, *CYP2C9*, *CYP4F2*, and *VKORC1* diplotypes (WGS Validation PGx group) were evaluated using the Geno4ME LDP PGx. Of these samples, six diplotype calls across five NHGRI Coriell samples could not be evaluated due to the diplotype being unknown or not supported by the Geno4ME LDP PGx pipeline (Supplemental Spreadsheet F). For diplotypes supported by the Geno4ME LDP PGx, the overall concordance between the Geno4ME LDP PGx and the previously characterized NHGRI Coriell samples was 100% (Table 8).

**Table 8.** PGx variant concordance of the Geno4ME LDP method and DNA derived from previously characterized NHGRI Coriell samples.

| Cohort size | Number of matching samples | Gene | Concordant variants |
| --- | --- | --- | --- |
| N=18 | 18/18 (100%) | *CYP2C19* | 18/18 (100%) |
|  |  | *CYP2C9* | 16/16 (100%) |
|  |  | *CYP4F2* | 14/14 (100%) |
|  |  | *VKORC1* | 18/18 (100%) |

### Concordance of Geno4ME LDP PGx to ORM PGx

Performance of the Geno4ME LDP PGx was further evaluated by comparing Geno4ME LDP PGx results of the WGS Validation ORM validation group to the OS-ORM PGx (N=188). The most common diplotype for each gene was *CYP2C19* *1/*1, CYP2C9 *1/*1, CYP4F2 *1/*1, and *VKORC1* GA (Table 9). Most participants in the WGS Validation ORM validation group had normal metabolizer status for *CYP2C19* and *CYP2C9* (Table 10). Slightly more than half of participants were classified as warfarin resistant based on *CYP4F2* diplotype and/or were warfarin sensitive based on *VKORC1* diplotype. For one participant, gme-039, the *CYP2C19* diplotype was identified using the Geno4ME LDP method as *4A/*17 and the OS-ORM PGx as *4/*17. However, the discrepancy was determined to be due to nomenclature differences of reporting *4 suballeles where *4A and *4 have the same core alleles and phenotype. The overall concordance between the Geno4ME LDP and ORM PGx diplotyping for *CYP2C19*, *CYP2C9*, *CYP4F2*, and *VKORC1* was 100% (Table 11, Supplemental Spreadsheet G).

**Table 9.** Distribution of PGx gene diplotypes from the WGS validation cohort identified using the Geno4ME LDP.

| Gene | Diplotype | n | Percent |
| --- | --- | --- | --- |
| *CYP2C19* | *1/*1 | 73 | 38.8% |
|  | *1/*17 | 50 | 26.6% |
|  | *1/*2 | 35 | 18.6% |
|  | *2/*17 | 13 | 6.9% |
|  | *17/*17 | 8 | 4.3% |
|  | *2/*2 | 5 | 2.7% |
|  | *1/*3 | 1 | 0.5% |
|  | *2/*4B | 1 | 0.5% |
|  | *4A/*17 | 1 | 0.5% |
|  | *8/*17 | 1 | 0.5% |
| *CYP2C9* | *1/*1 | 127 | 67.6% |
|  | *1/*2 | 36 | 19.1% |
|  | *1/*3 | 20 | 10.6% |
|  | *2/*3 | 3 | 1.6% |
|  | *2/*2 | 2 | 1.1% |
| *CYP4F2* | *1/*1 | 88 | 46.8% |
|  | *1/*3 | 85 | 45.2% |
|  | *3/*3 | 15 | 8.0% |
| *VKORC1* | GA | 109 | 58.0% |
|  | GG | 61 | 32.4% |
|  | AA | 18 | 9.6% |
| rs12777823 | GG | 132 | 70.2% |
|  | GA | 53 | 28.2% |
|  | AA | 3 | 1.6% |

**Table 10.** PGx metabolizer status distribution from the WGS validation cohort.

| Gene | Metabolizer Status | n | Percent |
| --- | --- | --- | --- |
| *CYP2C19* | Normal metabolizer | 73 | 38.8% |
|  | Intermediate metabolizer | 51 | 27.1% |
|  | Rapid metabolizer | 50 | 26.6% |
|  | Ultrarapid metabolizer | 8 | 4.3% |
|  | Poor Metabolizer | 6 | 3.2% |
| *CYP2C9* | Normal metabolizer | 127 | 67.6% |
|  | Intermediate metabolizer | 56 | 29.8% |
|  | Poor metabolizer | 5 | 2.7% |
| *CYP4F2* | Warfarin resistant | 100 | 53.2% |
|  | Normal warfarin sensitivity | 88 | 46.8% |
| *VKORC1* | Warfarin sensitive | 127 | 67.6% |
|  | Normal warfarin sensitivity | 61 | 32.4% |
| rs12777823 | Normal warfarin sensitivity | 132 | 70.2% |
|  | Warfarin sensitive | 56 | 29.8% |

**Table 11.** Concordance of the Geno4ME-LDP method and the OS-ORM PGx for identifying PGx diplotypes in the WGS Validation cohort.

| Cohort | Experimental Method | Reference Method | Gene Tested | Number of concordant results | Percent Concordance |
| --- | --- | --- | --- | --- | --- |
| WGS Validation ORM | Geno4ME-LDP | MALDI-TOF Genotyping | *CYP2C19* | 188 | 100% |
|  |  |  | *CYP2C9* | 188 | 100% |
|  |  |  | *VKORC1* | 188 | 100% |
|  |  |  | *CYP4F2* | 188 | 100% |

### Concordance between Blood and Saliva samples using the Geno4ME LDP

Of the 60 participants with matched blood and saliva samples that were processed using the Geno4ME LDP (WGS Validation SB validation group), 32 matched pairs were positive for P/LP or VUS variants and 28 were screen-negative (Table 12). Within the 32 positive matched pairs, the majority of the 47 matching variants were missense (Figure 6A). These 47 variants were identified across 26 different genes and the overall concordance between DNA derived from blood or saliva was 100% (Figure 6B, Supplementary Spreadsheet H). For these same 60 participants, Geno4ME LDP PGx diplotype results for *CYP2C19*, *CYP2C9*, *CYP4F2*, *VKORC1*, and rs12777823 were compared and had an overall concordance of 100% (Table 13, Supplemental Spreadsheet I).

**Figure 6.**
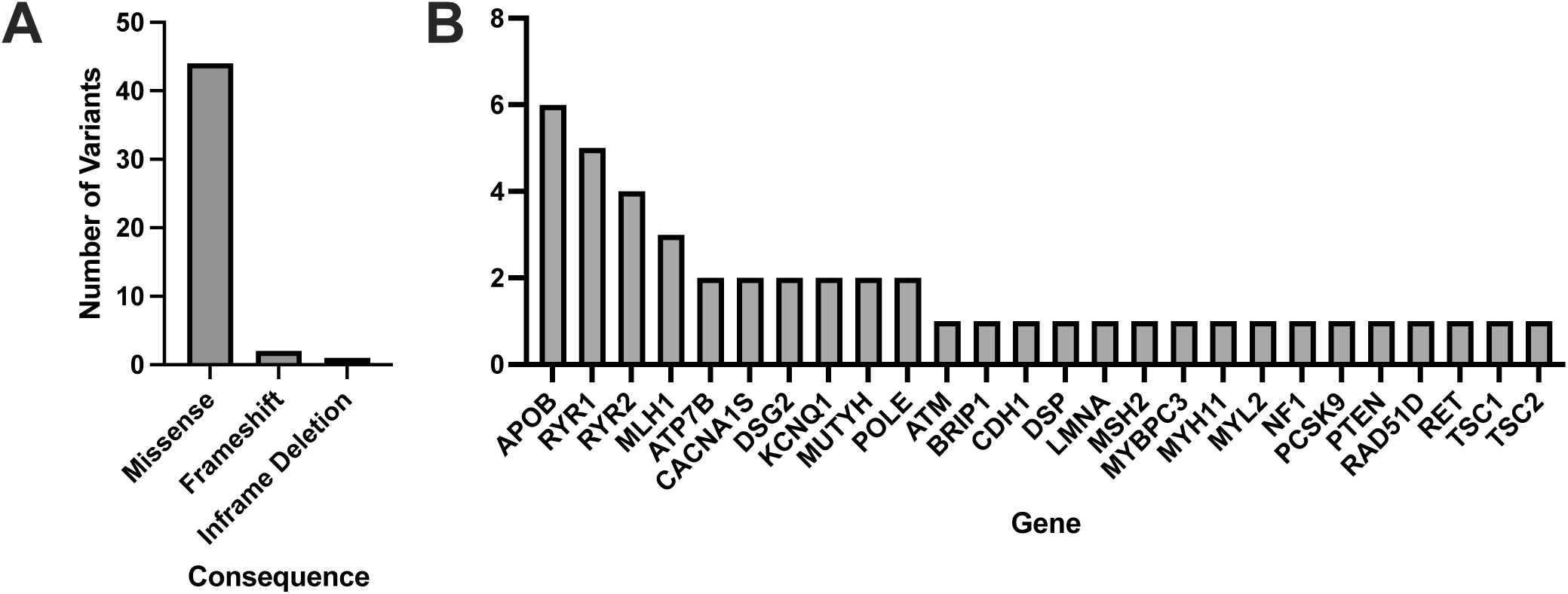
Molecular consequences and genes represented for all matched variants identified in the WGS Validation cohort comparing DNA derived from blood or saliva using Geno4ME LDP. A total of 47 variants were identified across all samples, with the majority being missense (A), and 26 unique genes were represented (B).

**Table 12.** Small variant concordance of the WGS-Fabric method between DNA derived from Blood or Saliva for the same participants.

| Cohort | Outcome | Number of matching samples | Concordant variants |
| --- | --- | --- | --- |
| WGS Validation SB | Positive | 32/32 (100%) | 47/47 (100%) |
|  | Negative | 28/28 (100%) | NA |

**Table 13.** PGx variant concordance of the Geno4ME LDP method between DNA derived from Blood or Saliva for the same participants.

| Cohort | Number of matching samples | Gene | Concordant variants |
| --- | --- | --- | --- |
| WGS Validation SB | 60/60 (100%) | *CYP2C19* | 60/60 (100%) |
|  |  | *CYP2C9* | 60/60 (100%) |
|  |  | *CYP4F2* | 60/60 (100%) |
|  |  | *VKORC1* | 60/60 (100%) |
|  |  | rs12777823 | 60/60 (100%) |

## Discussion

Rapid increases in sequencer capacity and throughput, automation and AI assistance for variant detection and interpretation, and significant decreases in per base sequencing costs are making it feasible to use WGS for routine clinical applications. Given that a majority of individuals carrying an actionable P/LP variant are unaware of their carrier status [56], and that accumulating evidence suggests that identifying individuals for testing based on medical history alone insufficiently captures the majority of individuals with monogenic risk for a heritable genetic condition [1, 2, 57], the introduction of WGS-based screening of patient populations has high potential for improving health outcomes. Furthermore, several recent studies have demonstrated the utility of population-based high-throughput genomic screening for facilitating early disease detection, risk management, and reducing adverse drug reactions [58–61].

To address this opportunity for improving health outcomes, we show that the Geno4ME LDP is a highly accurate clinical procedure for evaluating actionable genomic variants. We found 100% of previously tested variants obtained from patient EHRs to be identified using the DRAGEN WGS variant caller. The Geno4ME LDP had high overall agreement with well-characterized reference samples and with samples orthogonally tested by the ORM or WES-RM. When compared to variants identified using the OS-ORM PGx, the Geno4ME LDP had 100% sensitivity and specificity for identifying variants of interest. In addition, the Geno4ME LDP had 100% accuracy when compared to CNV and PGx reference samples. One notable discrepancy was the presence of a predicted pathogenic *TP53* c.854A>T p.Glu285Val in one WES sample but absent using Geno4ME LDP. The low level VAF of the *TP53* variant in the matched tumor sample compared to the control blood sample suggested that the variant may not be germline, but potentially clonal hematopoiesis of indeterminate potential (CHIP). However, the patient was deceased at the time of Geno4ME LDP testing and therefore additional sample could not be obtained to definitively determine the origin of the *TP53* variant in the WES data. This discrepancy demonstrates the need for genomic screening clinical tests to be understood in the context of the patient’s/participant’s medical history, and like all germline testing the ability to assess fibroblast cell lines if needed, particularly with TP53 variants.

The feasibility of using a clinical lab procedure such as Geno4ME LDP for large populations requires scalability at both the levels of sample collection and data analysis. We found identical variant concordance between samples obtained using saliva or blood. Our work agrees with previous data suggesting that once oral microbiome reads are excluded, saliva-derived and blood-derived genomic DNA are generally comparable in quality for clinical WGS workflows [62, 63]. Since saliva can be obtained via at-home collections without the need for a phlebotomist, the sample type would be more conducive to population health applications. At the level of data analysis, use of ACE in the variant curation process rapidly screened variants of interest and allowed for reduced hands-on time for manually curating variants. For example, out of 113,096 evaluated variants in the WGS Validation ORM group heritable disorder gene panel, ACE was able to reduce the number of variants requiring manual evaluation to 250 (0.02% of all variants evaluated in the group) without loss of P/LP identification sensitivity. In addition, the automated Geno4ME LDP PGx accurately diplotyped all samples, providing predicted metabolizer status for all PGx genes analyzed in this study without manual intervention.

Upfront testing of the whole genome using a method such as Geno4ME LDP allows for simultaneous testing of multiple genomic features while also allowing for future incorporation of additional screened genes and/or polygenic risk tests through region unmasking. Because the Geno4ME LDP was validated for all variant types of interest, new genes can rapidly be added to the panel in the future. Yet, there remains challenges when using short-read sequencing for achieving comprehensive population genomics testing. Recent work suggests that complex structural variants represent a substantial component of actionable genomic conditions [64]. In addition, highly polymorphic genes such as relevant PGx gene *CYP2D6*, pseudogene regions, and highly repetitive regions continue to present analysis challenges [65–67]. Future comparisons of the Geno4ME LDP to well-characterized complex reference samples will determine the limits of the method for detecting actionable SV and polymorphic variation as part of clinical testing.

## Data Availability

The raw datasets generated and/or analyzed during the current study are not publicly available due to protocol and Informed Consent language that mandate of IRB to review projects and require a data use agreement (DUA), but are available from the corresponding author on reasonable request. Analyzed data is contained within the manuscript and supplementary information files.

