## Supplementary Material for "Design and validation of a clinical whole genome sequencing-based assay for patient screening in a large healthcare system"

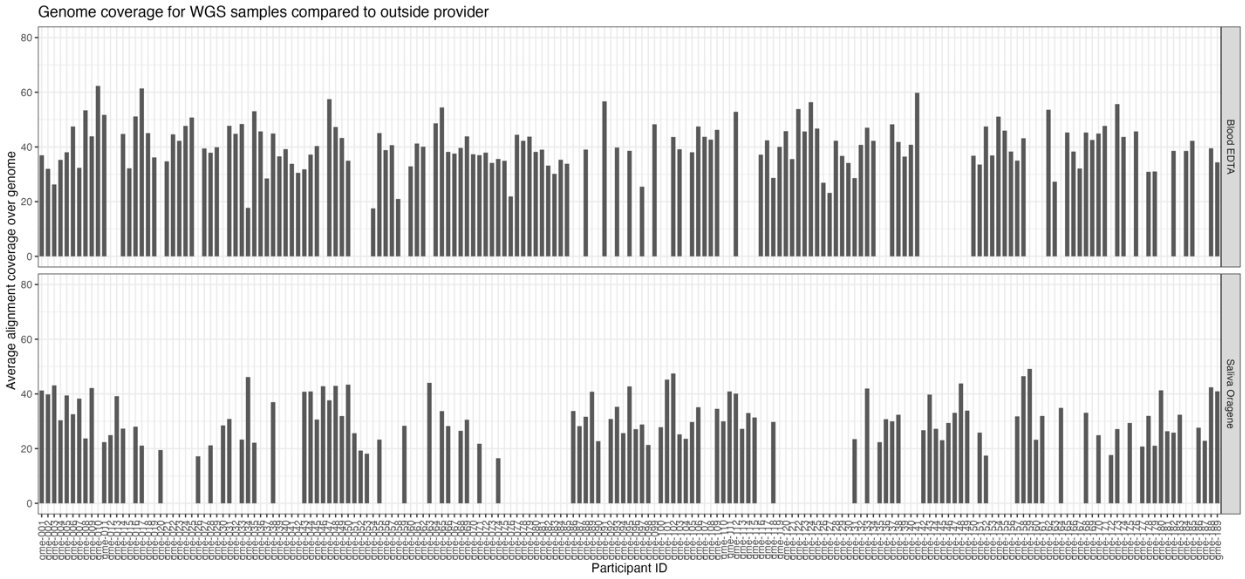


Figure S1 Average alignment coverage over genome for WGS samples in the WGS Validation cohort.

Figure S2. Average alignment coverage over genome for PGx, WES Comparison, and WGS CNV Validation cohorts.

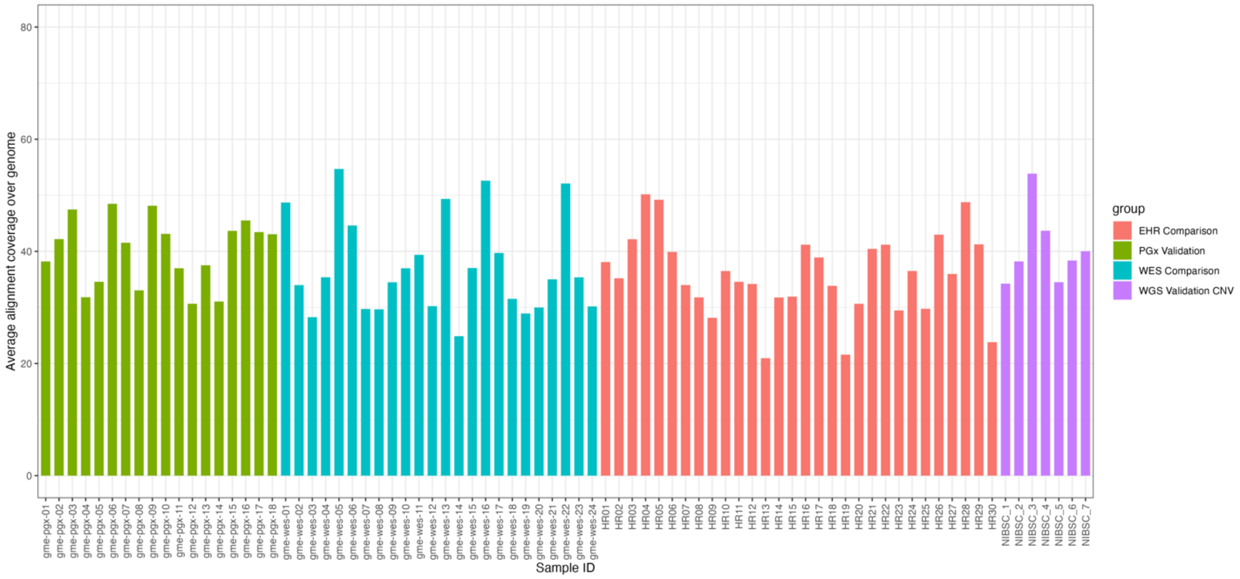

Figure S3. Average alignment coverage over genome (hg19) by method validation group.

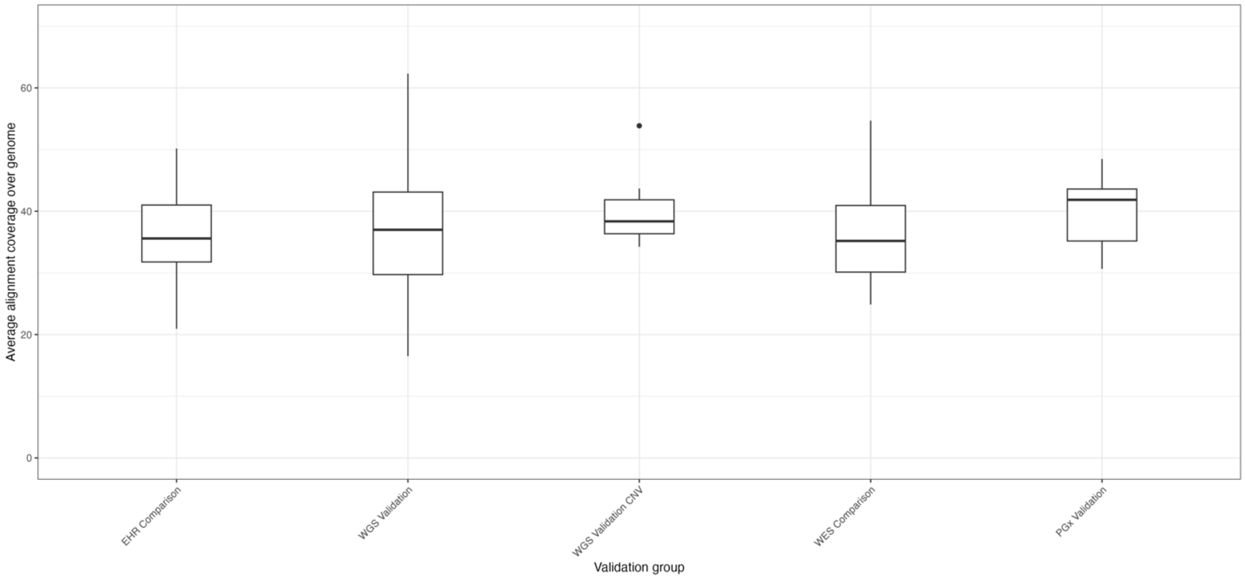

Figure S4. Total number of variants per case that was interpreted by Fabric in the WGS validation cohort.

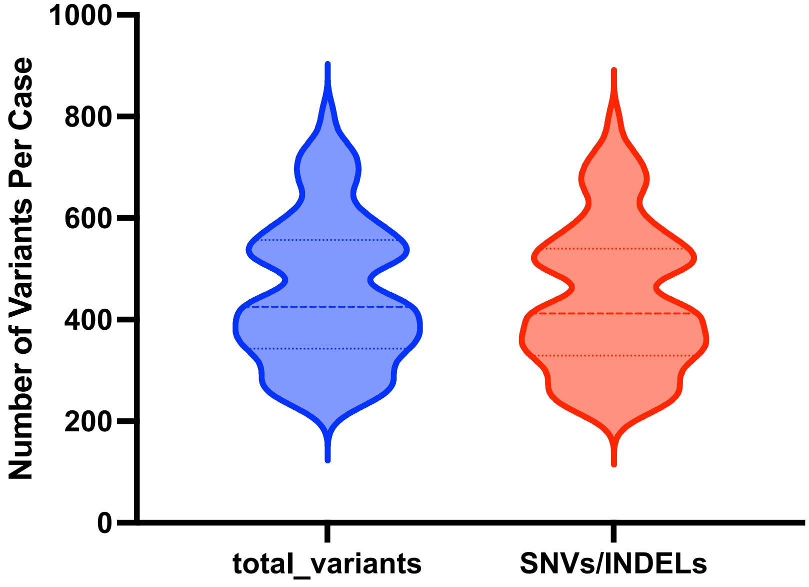

Figure S5. Sensitivity, Specificity calculations for WGS-Fabric method using outside reference method (ORM).

| | | OUTSIDE REFERENCE METHOD GENE VARIANT DETECTION | |
|---|---|---|---|
| | | + | - |
| WGS-FM GENE VARIANT DETECTION | + | 17 | 0 |
| | - | 0 | 14382 |

| ANALYTIC SPECIFICITY | |
|---|---|
| True Negative / (False Positive + True Negative) | |
| True Negative Total | 14382 |
| False Positive Total | 0 |
| False Positive + True Negative | 14382 |
| **Analytical Specificity** | **100.00%** |
| **95% CI** | **99.97 - 100.00%** |

| ANALYTIC SENSITIVITY | |
|---|---|
| True Positive / (True Positive + False Negative) | |
| True Positive Total | 17 |
| False Negative Total | 0 |
| True Positive + False Negative | 17 |
| **Analytical Sensitivity** | **100.00%** |
| **95% CI** | **80.49 - 100%** |

| TECHNICAL ACCURACY | |
|---|---|
| True Positive + True Negative / (Total Tested) | |
| True Positive Total | 17 |
| True Negative Total | 14382 |
| Total Tested | 14399 |
| **Technical Accuracy** | **100.00%** |
| **95% CI** | **99.97 - 100.00%** |

| NEGATIVE PREDICTIVE VALUE | |
|---|---|
| True Negative / (True Negative + False Negative) | |
| True Negative Total | 14382 |
| False Negative Total | 0 |
| True Negative + False Negative | 14382 |
| **Negative Predictive Value** | **100.00%** |
| **95% CI** | **99.97 - 100.00%** |

| POSITIVE PREDICTIVE VALUE | |
|---|---|
| True Positive / (True Positive + False Positive) | |
| True Positive Total | 17 |
| False Positive Total | 0 |
| True Positive + False Positive | 17 |
| **Positive Predictive Value** | **100.00%** |
| **95% CI** | **80.49 - 100%** |

Figure S6. Sensitivity, Specificity calculations for WGS-Fabric method using WES-ORM.

| | | REFERENCE ASSAY GENE VARIANT DETECTION | |
|---|---|---|---|
| | | + | - |
| WGS-FM GENE VARIANT DETECTION | + | 77 | 0 |
| | - | 1 | 874 |

| ANALYTIC SPECIFICITY<br>True Negative /<br>(False Positive + True Negative) | |
|---|---|
| True Negative Total | 874 |
| False Positive Total | 0 |
| False Positive + True Negative | 874 |
| **Analytical Specificity** | **100.00%** |
| **95% CI** | **99.58 - 100.00%** |

| ANALYTIC SENSITIVITY<br>True Positive /<br>(True Positive + False Negative) | |
|---|---|
| True Positive Total | 77 |
| False Negative Total | 1 |
| True Positive + False Negative | 78 |
| **Analytical Sensitivity** | **98.72%** |
| **95% CI** | **93.06 - 99.97%** |

| TECHNICAL ACCURACY<br>True Positive + True Negative /<br>(Total Tested) | |
|---|---|
| True Positive Total | 77 |
| True Negative Total | 874 |
| Total Tested | 952 |
| **Technical Accuracy** | **99.89%** |
| **95% CI** | **99.42 - 100.00%** |

| NEGATIVE PREDICTIVE VALUE<br>True Negative /<br>(True Negative + False Negative) | |
|---|---|
| True Negative Total | 874 |
| False Negative Total | 1 |
| True Negative + False Negative | 875 |
| **Negative Predictive Value** | **99.89%** |
| **95% CI** | **99.2 - 99.98%** |

| POSITIVE PREDICTIVE VALUE<br>True Positive /<br>(True Positive + False Positive) | |
|---|---|
| True Positive Total | 77 |
| False Positive Total | 0 |
| True Positive + False Positive | 77 |
| **Positive Predictive Value** | **100.00%** |
| **95% CI** | **95.32- 100.00%** |

Supplemental Table S1. Outside reference method gene panels (OS-ORM) for genomic conditions surveyed in this study

| **Gene** | **Panel** | **VUS reported** |
|---|---|---|
| ACTA2 | ORM Panel A | no |
| ACTC1 | ORM Panel A | no |
| APC | ORM Panel A | no |
| APOB | ORM Panel A | no |
| ATP7B | ORM Panel A | no |
| BMPR1A | ORM Panel A | no |
| BRCA1 | ORM Panel A | no |
| BRCA2 | ORM Panel A | no |
| CACNA1S | ORM Panel A | no |
| COL3A1 | ORM Panel A | no |
| DSC2 | ORM Panel A | no |
| DSG2 | ORM Panel A | no |
| DSP | ORM Panel A | no |
| FBN1 | ORM Panel A | no |
| GLA | ORM Panel A | no |
| KCNH2 | ORM Panel A | no |
| KCNQ1 | ORM Panel A | no |
| LDLR | ORM Panel A | no |
| LMNA | ORM Panel A | no |
| MEN1 | ORM Panel A | no |
| MLH1 | ORM Panel A | no |
| MSH2 | ORM Panel A | no |
| MSH6 | ORM Panel A | no |
| MUTYH | ORM Panel A | no |
| MYBPC3 | ORM Panel A | no |
| MYH11 | ORM Panel A | no |
| MYH7 | ORM Panel A | no |
| MYL2 | ORM Panel A | no |
| MYL3 | ORM Panel A | no |
| NF2 | ORM Panel A | no |
| OTC | ORM Panel A | no |
| PCSK9 | ORM Panel A | no |

| | | |
|---|---|---|
| PKP2 | ORM Panel A | no |
| PMS2 | ORM Panel A | no |
| PRKAG2 | ORM Panel A | no |
| PTEN | ORM Panel A | no |
| RB1 | ORM Panel A | no |
| RET | ORM Panel A | no |
| RYR1 | ORM Panel A | no |
| RYR2 | ORM Panel A | no |
| SCN5A | ORM Panel A | no |
| SDHAF2 | ORM Panel A | no |
| SDHB | ORM Panel A | no |
| SDHC | ORM Panel A | no |
| SMAD3 | ORM Panel A | no |
| SMAD4 | ORM Panel A | no |
| STK11 | ORM Panel A | no |
| TGFBR1 | ORM Panel A | no |
| TGFBR2 | ORM Panel A | no |
| TMEM43 | ORM Panel A | no |
| TNNI3 | ORM Panel A | no |
| TNNT2 | ORM Panel A | no |
| TP53 | ORM Panel A | no |
| TPM1 | ORM Panel A | no |
| TSC1 | ORM Panel A | no |
| TSC2 | ORM Panel A | no |
| VHL | ORM Panel A | no |
| WT1 | ORM Panel A | no |
| APC | ORM Panel B | yes |
| ATM | ORM Panel B | yes |
| AXIN2 | ORM Panel B | yes |
| BMPR1A | ORM Panel B | yes |
| BRCA1 | ORM Panel B | yes |
| BRCA2 | ORM Panel B | yes |
| BRIP1 | ORM Panel B | yes |
| CDH1 | ORM Panel B | yes |
| CDK4 | ORM Panel B | yes |
| CDKN2A | ORM Panel B | yes |
| CHEK2 | ORM Panel B | yes |
| EPCAM | ORM Panel B | yes |
| GREM1 | ORM Panel B | yes |
| MEN1 | ORM Panel B | yes |

| | | |
|---|---|---|
| MLH1 | ORM Panel B | yes |
| MSH2 | ORM Panel B | yes |
| MSH3 | ORM Panel B | yes |
| MSH6 | ORM Panel B | yes |
| MUTYH | ORM Panel B | yes |
| NBN | ORM Panel B | yes |
| NF1 | ORM Panel B | yes |
| NF2 | ORM Panel B | yes |
| NTHL1 | ORM Panel B | yes |
| PALB2 | ORM Panel B | yes |
| PMS2 | ORM Panel B | yes |
| POLD1 | ORM Panel B | yes |
| POLE | ORM Panel B | yes |
| PTEN | ORM Panel B | yes |
| RAD51C | ORM Panel B | yes |
| RAD51D | ORM Panel B | yes |
| RB1 | ORM Panel B | yes |
| RET | ORM Panel B | yes |
| SDHAF2 | ORM Panel B | yes |
| SDHB | ORM Panel B | yes |
| SDHC | ORM Panel B | yes |
| SDHD | ORM Panel B | yes |
| SMAD4 | ORM Panel B | yes |
| STK11 | ORM Panel B | yes |
| TP53 | ORM Panel B | yes |
| TSC1 | ORM Panel B | yes |
| TSC2 | ORM Panel B | yes |
| VHL | ORM Panel B | yes |
| WT1 | ORM Panel B | yes |
| CYP2C19 | ORM PGx | no |
| CYP2C9 | ORM PGx | no |
| CYP4F2 | ORM PGx | no |
| VKORC1 | ORM PGx | no |

Table S2. Refseq transcripts used by the WGS-FM.

| Gene | Transcript |
|---|---|
| ACTA2 | NM_001613.2 |
| ACTC1 | NM_005159.4 |
| APC | NM_000038.5 |
| APOB | NM_000384.2 |
| ATM | NM_000051.3 |
| ATP7B | NM_000053.3 |
| AXIN2 | NM_004655.3 |
| BMPR1A | NM_004329.2 |
| BRCA1 | NM_007294.3 |
| BRCA2 | NM_000059.3 |
| BRIP1 | NM_032043.2 |
| CACNA1S | NM_000069.2 |
| CDH1 | NM_004360.3 |
| CDK4 | NM_000075.3 |
| CDKN2A | NM_000077.4 |
| CHEK2 | NM_007194.3 |
| COL3A1 | NM_000090.3 |
| DSC2 | NM_024422.3 |
| DSG2 | NM_001943.3 |
| DSP | NM_004415.2 |
| EPCAM | NM_002354.2 |
| FBN1 | NM_000138.4 |
| GLA | NM_000169.2 |
| GREM1 | NM_013372.6 |
| KCNH2 | NM_000238.3 |
| KCNQ1 | NM_000218.2 |
| LDLR | NM_000527.4 |
| LMNA | NM_170707.3 |
| MEN1 | NM_130799.2 |
| MLH1 | NM_000249.3 |
| MSH2 | NM_000251.2 |
| MSH3 | NM_002439.4 |
| MSH6 | NM_000179.2 |
| MUTYH | NM_001128425.1 |
| MYBPC3 | NM_000256.3 |
| MYH11 | NM_002474.2 |
| MYH7 | NM_000257.2 |

| | |
|---|---|
| MYL2 | NM_000432.3 |
| MYL3 | NM_000258.2 |
| NBN | NM_002485.4 |
| NF1 | NM_000267.3 |
| NF2 | NM_000268.3 |
| NTHL1 | NM_002528.5 |
| OTC | NM_000531.5 |
| PALB2 | NM_024675.3 |
| PCSK9 | NM_174936.3 |
| PKP2 | NM_004572.3 |
| PMS2 | NM_000535.5 |
| POLD1 | NM_002691.3 |
| POLE | NM_006231.2 |
| PRKAG2 | NM_016203.3 |
| PTEN | NM_000314.4 |
| RAD51C | NM_058216.2 |
| RAD51D | NM_002878.3 |
| RB1 | NM_000321.2 |
| RET | NM_020975.4 |
| RYR1 | NM_000540.2 |
| RYR2 | NM_001035.2 |
| SCN5A | NM_198056.2 |
| SDHAF2 | NM_017841.2 |
| SDHB | NM_003000.2 |
| SDHC | NM_003001.3 |
| SDHD | NM_003002.3 |
| SMAD3 | NM_005902.3 |
| SMAD4 | NM_005359.5 |
| STK11 | NM_000455.4 |
| TGFBR1 | NM_004612.2 |
| TGFBR2 | NM_003242.5 |
| TMEM43 | NM_024334.2 |
| TNNI3 | NM_000363.4 |
| TNNT2 | NM_001001430.2 |
| TP53 | NM_000546.5 |
| TPM1 | NM_001018005.1 |
| TSC1 | NM_000368.4 |
| TSC2 | NM_000548.3 |
| VHL | NM_000551.3 |
| WT1 | NM_024426.4 |

Table S3. Genotype to phenotype mapping for PGx based on PharmgKB, CPIC, and PharmVar annotations.

| Gene | Diplotypes/Genotypes | Phenotypes |
|---|---|---|
| CYP2C9 | Homozygote or compound heterozygote diplotype including *2, *3, *5, *6, *8, *11 | Poor metabolizer (PM) |
| | Heterozygote for *2, *3, *5, *6, *8, *11 | Intermediate metabolizer (IM) |
| | None of the tested alleles (*2, *3, *5, *6, *8, *11) detected. Reported as *1/*1 | Normal metabolizer (NM) |
| CYP2C19 | Homozygote or compound heterozygote diplotype including two no function alleles (*2, *3, *4A, *4B, *5, *6, *7, *8, *35) | Poor metabolizer (PM) |
| | Compound heterozygote diplotype including one decreased function allele (*9 or *10) and one no function allele (*2, *3, *4A, *4B, *5, *6, *7, *8, and *35) | Likely poor metabolizer (likely PM) |
| | Compound heterozygote diplotype including *17 or *1 (i.e. no *17 detected), and one no function allele (*2, *3, *4A, *4B, *5, *6, *7, *8, and *35) | Intermediate metabolizer (IM) |
| | Compound heterozygote diplotype including *17 or *1 (i.e. no *17 or other tested alleles detected), and one decreased function allele (*9 or *10)<br>OR<br>Homozygote or compound heterozygote diplotype including two decreased function alleles (*9 and *10) | Likely intermediate metabolizer (Likely IM) |
| | None of the tested alleles (*2, *3, *4A, *4B, *5, *6, *7, *8, *9, *10, *17, *35) detected. Reported as *1/*1 | Normal metabolizer (NM) |
| | Heterozygote for only *17 (no other tested alleles detected). Reported as *1/*17 | Rapid metabolizer (RM) |
| | Homozygote for *17 | Ultra rapid metabolizer (UM) |
| CYP4F2 | Heterozygote or homozygote for *3 | Warfarin resistant |
| | No *3 detected. Reported as *1/*1 | Normal warfarin sensitivity |
| VKORC1 (c.-1639G>A, rs9923231) | GA or AA | Warfarin sensitive |
| | GG | Normal warfarin sensitivity |
| | GA, AA | Warfarin sensitive |

| | | |
|---|---|---|
| rs12777823<br>(CYP2C gene cluster) | GG | Normal warfarin sensitivity |
